## Supplementary material for "The Contribution of Pharmacogenetic Drug Interactions to 90-Day Hospital Readmissions: Preliminary Results from a Real-World Healthcare System": https://docs.google.com/document/d/1t2fOr5XZi6HoA1rx23th2aWd56spuv4O/edit

**Supplemental Material**

**Table S1. List of Medication Orders/Administration from 12/01/2009 to 12/31/2020 with Orders 30 days Prior to Inpatient Admissions**

|  | **Readmission within 90 days of inpatient admission discharge date in Encounter Fact between 1/1/2010 and 3/31/2021 inclusive** | | |
| --- | --- | --- | --- |
| **CPIC medications** | **Overall** | **No** | **Yes** |
| **N (%)** | **2,211 (100)** | **1,935 (87.5)** | **276 (12.5)** |
| Amitriptyline | 26 (1.2) | 14 (0.7) | 12 (4.3) |
| Atazanavir | 0 (0) | 0 (0) | 0 (0) |
| Atomoxetine | 5 (0.2) | 3 (0.2) | 2 (0.7) |
| Azathioprine | 9 (0.4) | 5 (0.3) | 4 (1.4) |
| Capecitabine | 0 (0) | 0 (0) | 0 (0) |
| Celecoxib | 287 (13) | 238 (12.3) | 49 (17.8) |
| Citalopram | 24 (1.1) | 18 (0.9) | 6 (2.2) |
| Clomipramine | 2 (0.1) | 2 (0.1) | 0 (0) |
| Clopidogrel | 55 (2.5) | 36 (1.9) | 19 (6.9) |
| Codeine | 64 (2.9) | 45 (2.3) | 19 (6.9) |
| Desipramine | 0 (0) | 0 (0) | 0 (0) |
| Doxepin | 4 (0.2) | 0 (0) | 4 (1.4) |
| Efavirenz | 0 (0) | 0 (0) | 0 (0) |
| Escitalopram | 120 (5.4) | 93 (4.8) | 27 (9.8) |
| Fluorouracil | 4 (0.2) | 2 (0.1) | 2 (0.7) |
| Flurbiprofen | 0 (0) | 0 (0) | 0 (0) |
| Fluvoxamine | 1 (0) | 1 (0.1) | 0 (0) |
| Fosphenytoin | 4 (0.2) | 0 (0) | 4 (1.4) |
| Ibuprofen | 1064 (48.1) | 963 (49.8) | 101 (36.6) |
| Imipramine | 0 (0) | 0 (0) | 0 (0) |
| Lansoprazole | 31 (1.4) | 19 (1) | 12 (4.3) |
| Lornoxicam | 0 (0) | 0 (0) | 0 (0) |
| Meloxicam | 35 (1.6) | 29 (1.5) | 6 (2.2) |
| Mercaptopurine | 1 (0) | 1 (0.1) | 0 (0) |
| Nortriptyline | 20 (0.9) | 15 (0.8) | 5 (1.8) |
| Omeprazole | 541 (24.5) | 416 (21.5) | 125 (45.3) |
| Ondansetron | 2026 (91.6) | 1769 (91.4) | 257 (93.1) |
| Pantoprazole | 298 (13.5) | 204 (10.5) | 94 (34.1) |
| Paroxetine | 24 (1.1) | 16 (0.8) | 8 (2.9) |
| Peginterferon-Alfa-2a | 0 (0) | 0 (0) | 0 (0) |
| Peginterferon-Alfa-2b | 0 (0) | 0 (0) | 0 (0) |
| Phenytoin | 6 (0.3) | 3 (0.2) | 3 (1.1) |
| Piroxicam | 0 (0) | 0 (0) | 0 (0) |
| Sertraline | 102 (4.6) | 84 (4.3) | 18 (6.5) |
| Simvastatin | 91 (4.1) | 66 (3.4) | 25 (9.1) |
| Tacrolimus | 10 (0.5) | 5 (0.3) | 5 (1.8) |
| Tamoxifen | 6 (0.3) | 6 (0.3) | 0 (0) |
| Tenoxicam | 0 (0) | 0 (0) | 0 (0) |
| Thioguanine | 0 (0) | 0 (0) | 0 (0) |
| Tramadol | 347 (15.7) | 252 (13) | 95 (34.4) |
| Trimipramine | 0 (0) | 0 (0) | 0 (0) |
| Tropisetron | 0 (0) | 0 (0) | 0 (0) |
| Voriconazole | 0 (0) | 0 (0) | 0 (0) |
| Warfarin | 237 (10.7) | 185 (9.6) | 52 (18.8) |

### **Table S2. Frequency of 90-day Readmission Diagnostic Categories (N=281)**

| **Diagnostic Categories** | **Frequency** | **Percent** |
| --- | --- | --- |
| Behavioral/Psychiatric | 27 | 9.82 |
| Cancer/Neoplasm | 32 | 11.64 |
| Cardiovascular | 16 | 5.82 |
| Developmental/Disability | 1 | 0.36 |
| Endocrine/Metabolic | 48 | 17.45 |
| Gastrointestinal | 30 | 10.91 |
| Genitourinary | 3 | 1.09 |
| Gynecological | 1 | 0.36 |
| Hematological | 16 | 5.82 |
| Infection/Abscess | 2 | 0.73 |
| Infection/Abscess/Other | 9 | 3.27 |
| Infection/Dermatological/Abscess | 3 | 1.09 |
| Infection/Gastrointestinal | 9 | 3.27 |
| Infection/Genitourinary | 3 | 1.09 |
| Infection/Respiratory | 5 | 1.82 |
| Inflammatory/Rheumatological | 2 | 0.73 |
| Neurological | 10 | 3.64 |
| Obstetrical | 21 | 7.64 |
| Orthopedic/Musculoskeletal | 13 | 4.73 |
| Pain | 4 | 1.45 |
| Pulmonary | 10 | 3.64 |
| Vascular | 1 | 0.36 |
| Vascular/Non-Cardiac | 9 | 3.27 |

*6 patients missing readmission diagnosis.
